# Modelling the impact of climate variability on the effectiveness of seasonal indoor residual spraying

**DOI:** 10.64898/2026.01.31.26345292

**Authors:** Alexis Martin-Makowka, James D. Munday, Adrian Mark Tompkins, Cyril Caminade, Nakul Chitnis

## Abstract

Malaria transmission is strongly modulated by climate, yet most mechanistic models used for health policy evaluation do not explicitly account for climate-driven variability in mosquito dynamics. Here, we present a novel modelling framework that couples VECTRI, a climate-sensitive model for malaria, with OpenMalaria, a detailed individual-based model of malaria epidemiology and intervention impact. This integrated approach enables simulation of transmission processes that respond to interannual climate fluctuations while retaining the capacity to evaluate realistic intervention strategies.

We apply this framework to Mozambique, a high-burden country for malaria, with pronounced climatic seasonality and extensive routine surveillance data. Using district-level incidence time series from the ten highest-incidence districts, we validate the modelling framework and compare its performance with the standalone versions of VECTRI and OpenMalaria. The joint modelling framework reproduces malaria seasonality more accurately than the two single models, with improved timing of incidence peaks in 7 out of 10 districts and a closer representation of evolution of transmission throughout the season, as measured by the seasonality index, in 7 out of 10 districts.

We then use the coupled system to assess how climate-driven interannual variability affects the predicted effectiveness of indoor residual spraying, a key seasonal malaria control intervention in Mozambique. Integrating climate-driven interannual variability into OpenMalaria substantially impacts the modeled effectiveness of indoor residual spraying. The joint modelling framework increases the estimated protective effectiveness of indoor residual spraying in reducing incidence by up to 11% and delayed the optimal deployment window by two weeks.

Our results demonstrate that climate-informed mechanistic models can meaningfully alter estimates of intervention impact and improve the realism of malaria predictions. The joint OpenMalaria-VECTRI modelling framework provides a flexible tool for national malaria programs seeking to evaluate seasonal interventions under varying climatic scenarios.

**Author summary:** Malaria transmission in Mozambique changes from year to year because mosquito populations strongly depend on rainfall and temperature. However, most models used to guide malaria control planning do not fully capture these climate-driven fluctuations. We developed a new modelling approach that links two existing tools: VECTRI, which simulates climate-sensitive mosquito and malaria dynamics, and OpenMalaria, which simulates malaria infections and the effects of interventions.

We validated this joint modelling framework using routine surveillance malaria data from Mozambique and found that it better captures observed seasonal and interannual patterns than standalone models. We then used it to explore how climate variability influences the effectiveness of indoor residual spraying, an important seasonal malaria control strategy. Accounting for climate-driven mosquito dynamics reduced the predicted protective effectiveness of indoor residual spraying by 11% and shifted the optimal deployment timing by approximately two weeks. These results show that integrating climate information into malaria models can improve the accuracy and usefulness of intervention planning.

## Introduction

Malaria is one of the most consequential vector-borne infectious diseases worldwide, imposing a massive socio-economic and health burden on tropical and subtropical regions (WHO 2025; Ferrari et al. 2024). It remains one of the greatest public health threats in Mozambique, the country of focus here, with over 10 million cases reported annually since 2018 (Harp et al.2021), accounting for 29% of all deaths and 42% of deaths among children under five (PMI 2023).

A broad arsenal of malaria prevention tools is available, including insecticide-treated nets, indoor residual spraying, chemoprevention strategies such as seasonal malaria chemoprevention, and more recently, malaria vaccines (Hemingway et al.2016). Since the early 2000s, large-scale deployment of some of these interventions has contributed to substantial reductions in transmission and mortality (Bhatt et al. 2015). Nevertheless, malaria continues to have a large health impact, and after two decades of global progress, recent evidence points to a concerning stagnation and in some settings even a rebound in malaria burden (WHO 2025).

Multiple factors have contributed to this recent rebound, including growing insecticide and drug resistance. Moreover, increasing attention has also been drawn to the role of climate variability and extreme weather events in shaping malaria risk. The 2023 WHO World Malaria Report (WHO 2023) underlines the devastating impact of the 2022 floods in Pakistan, which led to an explosive malaria epidemic, with a five-fold increase in malaria cases reported in 2023. Climate influences malaria transmission through several biological pathways. Temperature regulates mosquito development, survival, biting rate, and the speed of parasite replication within the mosquito (Mordecai et al. 2019; Craig et al. 1999). Rainfall controls the availability and persistence of larval habitats and food resources, thereby driving mosquito population dynamics (Thomson et al. 2017). As a consequence, mosquito population dynamics and by extension malaria transmission risk strongly respond to climatic variations (Reiner et al. 2015).

Mechanistic malaria models can provide a valuable framework for understanding transmission processes and assessing intervention strategies. It is clear that in order to further progress in reducing malaria burden and work towards elimination, modelling tools are required that can assess the impacts of a wide and growing arsenal of intervention tools while accounting for climate seasonality and the broader context of climate interannual variability, extremes and long-term change. Such tools would allow planners to assess how climate anomalies have confounded or enhanced past intervention effectiveness for example. Moreover, many interventions are most effective when timed to coincide with peak transmission periods (Wagman et al. 2020; Cairns et al. 2012). One example is indoor residual spraying, the protective effect of which typically lasts three to six months (Sahu et al. 2020; Coleman et al. 2017). Modelling tools that account for both climate and interventions could identify the most effective intervention timing according to the climate setting, possibly even permitted year-to-year honing of strategy if reliable forecasts allow. Aligning intervention timing with climatic drivers therefore remains a key challenge for national malaria control programs (NMCPs).

To date, modelling tools that account for climate in a comprehensive manner and incorporate extensively validated interventions do not exist. Several malaria transmission models incorporate climatic drivers to varying extents. The Liverpool Malaria Model explicitly represents the effects of temperature and precipitation on vector ecology and transmission dynamics, but it is primarily designed as a climate-driven transmission model and offers limited capacity for evaluating detailed intervention scenarios and does not account for population density (Ermert et al. 2011a; Ermert et al. 2011b; Hoshen and Morse 2004).

Above all, the Liverpool model is not open source, which is essential to ensure reproducibility, algorithmic transparency, and community-driven validation, all necessary to secure the public health trust. EMOD provides a framework for modelling malaria interventions. It includes climate inputs, but its integration is not yet usable for detailed climate-impact assessments (Eckhoff 2013; Eckhoff 2012; Eckhoff 2011). Finally, malariasimulation is a lightweight, compartmental modelling framework in which climatic variables can modulate seasonal forcing or transmission intensity. However, climate effects are not represented mechanistically through explicit simulation of vector ecology or life stages (Charles et al. 2025).

Two other models also exist that respectively address the climate and intervention modelling aspects rigorously, and have been fully open source since their inception and are thus widely adopted by the modelling community. VECTRI is a climate-driven malaria model that explicitly represents mosquito and parasite development as functions of temperature and rainfall, enabling detailed simulation of climate-transmission relationships (Tompkins and Ermert 2013)). OpenMalaria is a flexible individual-based model designed to evaluate a wide range of malaria interventions and their epidemiological effects in diverse settings (OpenMalaria 2026; Chitnis et al. 2012; Smith et al. 2012; Smith et al. 2008).

Since none of these models simultaneously offer a fully mechanistic representation of climate-driven vector dynamics together with a detailed and flexible framework for intervention effectiveness analysis, in this study, we introduce a new approach that couples the strengths of existing malaria models. We accomplish this by integrating VECTRI’s representation of climate-sensitive entomological processes with OpenMalaria’s capacity to simulate detailed intervention scenarios. This joint modelling framework allows to examine how climate variability affects both malaria transmission and the impact of interventions. We apply this coupled modelling approach to Mozambique, a country with a persistently high malaria burden and a well-established relationship between climate and malaria incidence (Armando et al. 2025).

A high-quality dataset from the National Malaria Control Program of Mozambique is available (Harp et al. 2021), enabling us to validate our framework for high-burden districts and assess the added value of integrating climate-driven and intervention-driven mechanistic models. This new framework allows us to incorporate climate-driven interannual variations in malaria transmission into a modelling tool designed for policy support (OpenMalaria), and we evaluate how accounting for these variations influences the predicted effectiveness of a standard seasonal intervention method: indoor residual spraying.

## Methods

### OpenMalaria

The modelling framework we describe in this paper is designed to account for interventions, with OpenMalaria responsible for capturing their effects. We use OpenMalaria version 47. OpenMalaria is a detailed individual-based stochastic simulation model designed to support malaria control policy by simulating *Plasmodium falciparum* transmission and the impact of interventions (OpenMalaria 2026; Chitnis et al. 2012; Smith et al. 2012; Smith et al. 2008). OpenMalaria consists of a human host module and a mosquito vector module that are interconnected. The human module simulates the within-host dynamics of malaria infections using an agent-based approach. When bitten by an infectious mosquito, a human host may become infected. The module tracks individual infection histories, including parasite densities, clinical symptoms, and the development of immunity over time. Host heterogeneity is represented explicitly, accounting for factors such as age, immunity status, and attractiveness to mosquitoes. The simulation proceeds in 5-day increments (pentads), updating each individual’s infection status and immunity profile. At each time step, the model outputs which individuals are infected, allowing aggregation into population-level metrics such as malaria incidence and prevalence.

The mosquito module is a nested compartmental model updated daily. It tracks the number of blood-seeking female mosquitoes and their infection status: any infection status (including susceptible mosquitoes), infected or infectious, or infectious only. A blood-seeking female mosquito can find a host or die while doing so. Once a mosquito successfully finds a host, it initiates a gonotrophic cycle that involves blood feeding, resting and ovipositing, before a subsequent host-seeking. The probability of a mosquito successfully finding a host depends on host availability and species-specific feeding preferences. At each step of the gonotrophic cycle, mosquitoes face a risk of mortality. If a mosquito feeds on an infectious human, it becomes infected and undergoes a sporogonic cycle, after which it becomes infectious. The gonotrophic cycles of *Anopheles spp*. is described in Figure 1.

**Figure 1:**
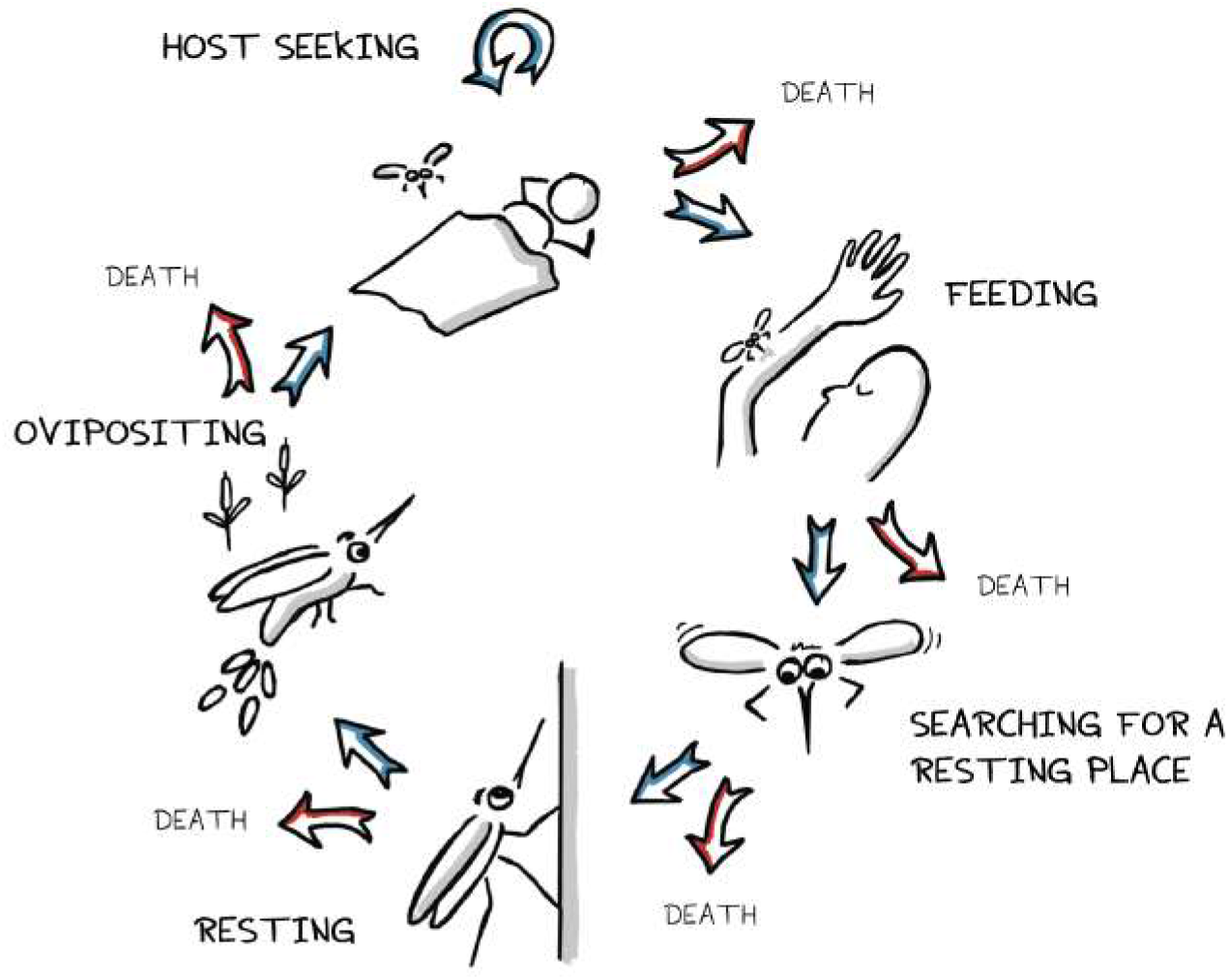
Gonotrophic cycle of *Anopheles spp*. mosquitoes. At each step of the gonotrophic cycle, female *Anopheles* mosquitoes can either reach the next step or die, with some probability modeled by OpenMalaria.

Simulations begin with a fully susceptible human population. Initially, only the human module is active, and malaria transmission is forced into the human population via an externally defined, strictly seasonal entomological inoculation rate (EIR) time series. This period during which malaria is forced into the human population lasts 90 years and allows for the buildup of immunity across a full generational turnover. After this period, the mosquito module is activated, and humans can now infect mosquitoes, which in turn can infect humans through a simulated EIR. However, the EIR driving malaria in humans is still guided by the input time series, and the model runs for an additional 30 years. During this phase, the emergence rate of mosquitoes is calibrated to ensure that the simulated EIR matches the input EIR. Once these two EIR time series are close enough, the input EIR can be removed, and the model proceeds autonomously with full vector-host dynamics for another 30-year burn-in period to stabilize malaria transmission. Following the burn-in period, interventions can be deployed to evaluate their effects on malaria transmission. OpenMalaria allows for parameter modifications representing diverse control measures, such as indoor residual spraying. Model outputs include the simulated EIR, clinical episode counts, and infection prevalence. Importantly, OpenMalaria does not explicitly model climatic drivers of transmission. Instead, seasonal variations must be incorporated by prescribing seasonal input EIR accordingly. These variations will be reflected in the computed emergence rate. A detailed description of model mechanics is available in Chitnis et al. (2012), Smith et al. (2012), and Smith et al. (2008) and on the OpenMalaria GitHub repository (OpenMalaria 2026). Figure 2 summarizes how the human and mosquito modules interact.

**Figure 2:**
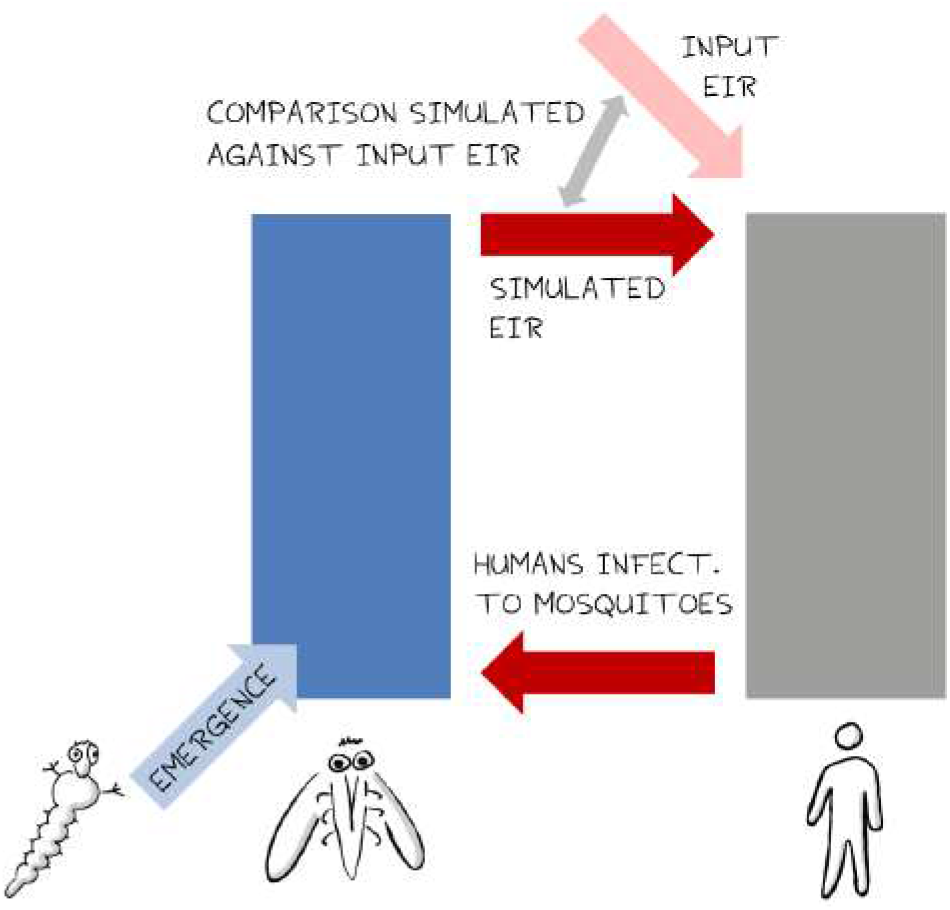
Overview of the human and mosquito modules of OpenMalaria and and their interaction. OpenMalaria consists of two interconnected modules: an individual-based human host module and a compartmental mosquito vector module. Initially, malaria transmission is forced using an external input entomological inoculation rate (EIR) to allow immunity to build up in the human population. The mosquito module is then introduced, enabling full vector-host transmission dynamics. The system runs forward with the possibility to include interventions, which can be applied flexibly as parameter modifications.

### VECTRI

The VECTRI model captures the effect of climate in the two-model framework. VECTRI (version 1.8.1) is a spatially explicit, climate-driven malaria model developed to simulate the impact of environmental factors on malaria dynamics at regional scales (Tompkins and Ermert 2013; Asare et al. 2016; Garrido Zornoza et al. 2024). Specifically designed for *Anopheles gambiae* and *Plasmodium falciparum*, the model incorporates the effects of temperature, rainfall, and human population density on vector and parasite development. VECTRI can operate on a grid-based spatial domain or can be applied to a single location. VECTRI simulates mosquito population dynamics through a bin-resolved compartmental structure representing the full aquatic and adult life stages, as shown in the schematic of the model components and transmission cycle (Figure 3). Larval development is influenced by water temperature and resource availability in breeding sites, which are modeled via a pond model forced by daily rainfall and air temperature data Asare et al. 2016. Eggs laid by adult mosquitoes enter the larval stage and progress through successive bins, facing a probability of death or maturation to pupae. Once emerging as adults, mosquitoes initiate gonotrophic cycles that include host seeking, blood feeding, resting and egg laying. Each stage is temperature-dependent and subject to mortality rates. Transmission is modeled dynamically: mosquitoes may acquire infection upon biting an infectious human, initiating a sporogonic cycle that is also bin-resolved and sensitive to ambient temperature. Infectious mosquitoes, in turn, can transmit malaria to susceptible humans, triggering a within-host development period (sporogonic cycle in humans). While mosquito parasite development is climate-dependent, human infection progression is modeled using a fixed developmental timeline.

**Figure 3:**
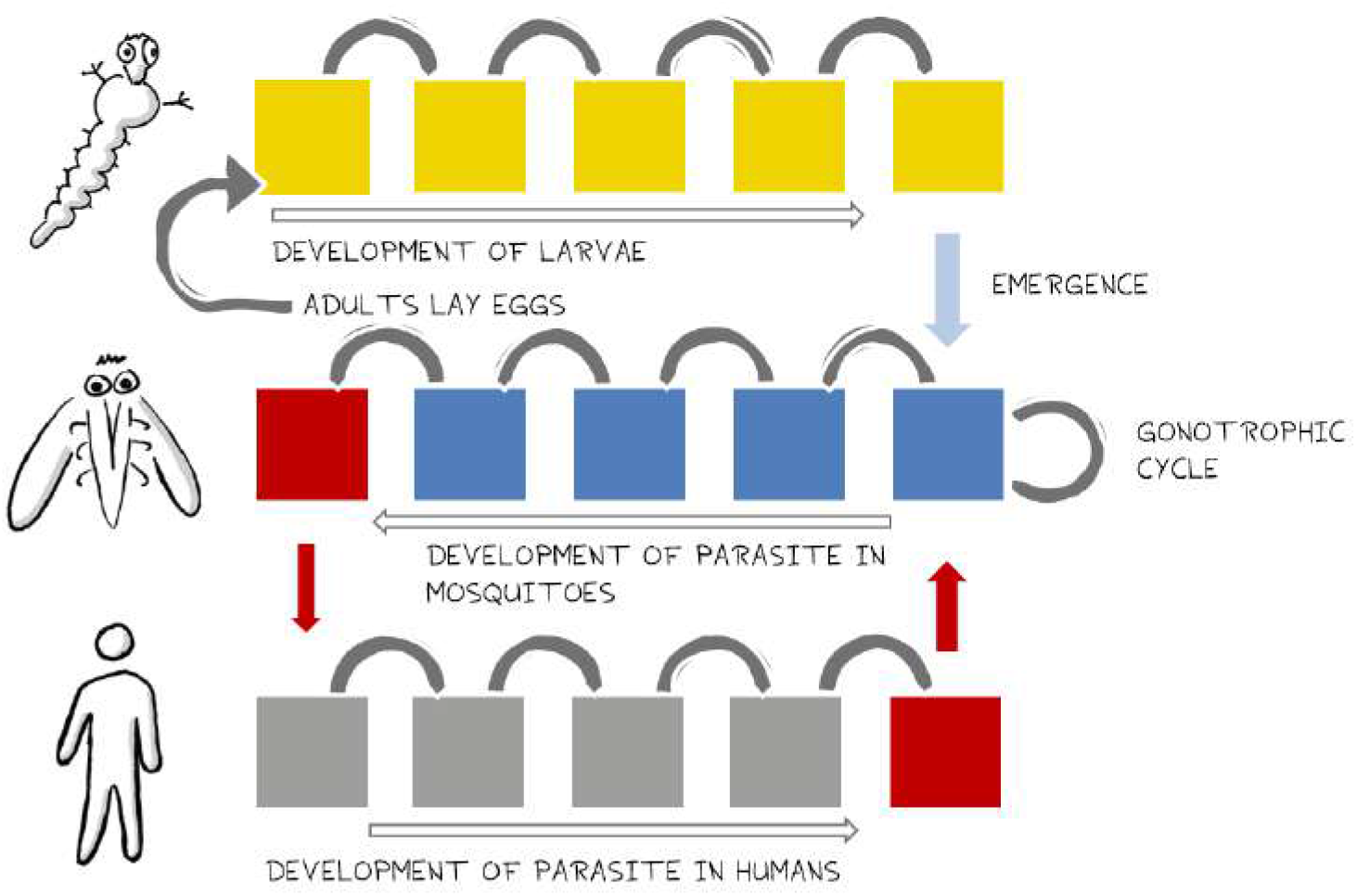
An overview of the VECTRI model. VECTRI consists of development bins tracking the progression of mosquito larvae and parasite development in mosquitoes and humans. The adult mosquito gonotrophic cycle is continuously resolved.

VECTRI runs on a daily timestep and includes a one-year spin-up period, starting from an initial proportion of infected humans to allow the system to reach equilibrium. Outputs include key entomological indicators such as mosquito emergence rate, adult vector density, and the EIR, as well as epidemiological metrics like parasite prevalence and malaria case incidence.

### Coupling of VECTRI with OpenMalaria

Our modeling framework integrates the climate-sensitive vector dynamics simulated by VECTRI into the transmission modeling environment of OpenMalaria. VECTRI simulations were conducted at a spatial resolution of 0.5°× 0.5° (approximately 50km × 50km) across the African continent, using daily air temperature at 2 meters and precipitation data from the CPC Global Unified Gauge-Based Analysis dataset for the period 1990–2023 (NOAA Physical Sciences Laboratory 2026). VECTRI also requires human population density data as input, here derived from the NASA’s Gridded Population of the World, version 4 (GPWv4) from CIESIN (2026), available at 1km spatial resolution and interpolated to match the climate data grid. From the VECTRI output, we use the daily mosquito emergence rate as an input to OpenMalaria. The daily emergence rate is aggregated to match the pentad (5-day) resolution used in OpenMalaria. We additionally record the mean annual EIR from VECTRI.

OpenMalaria simulation parameters are taken from the standard XML schema 47 (OpenMalaria 2024). Namely, population size is set to 10,000. Simulated mosquitoes are all *Anopheles gambiae*. The simulation, after initialization period, spans from January 1st, 2017 to August 31st, 2023.

OpenMalaria is initialized with a constant (flat) input EIR. The annual transmission level is set up to 10% of VECTRI’s mean annual EIR for the selected location, to account for the reduction of intensity resulting of unknown local interventions; the exact level of intensity at this stage is not crucial as we will recalibrate the malaria metrics later. During the burn-in period, the simulated EIR is still perennial and the mosquito emergence rate is kept constant. To incorporate the influence of VECTRI’s climate-driven mosquito dynamics, we adjust the mosquito emergence rate in OpenMalaria to follow the seasonal and interannual variability in the mosquito emergence rate simulated by VECTRI. This is implemented as a larval control intervention in OpenMalaria, where the constant emergence rate is modulated based on VECTRI’s relative deviations from its mean. Since the two models differ in scale (VECTRI expresses emergence per unit area while OpenMalaria uses absolute mosquito counts), we apply a relative scaling approach: the OpenMalaria emergence rate is adjusted proportionally to VECTRI’s deviation from its overall average. For example, if VECTRI’s emergence rate at a given time is half its 2017-2023 average, the emergence rate in OpenMalaria is also halved.

### Validation and comparison of modelling approaches

We validate the two-model framework using empirical malaria incidence data from Mozambique. In this country, transmission intensity varies substantially between regions and between years, driven by climatic and socioeconomic factors. In Mozambique, *Anopheles funestus sensu stricto* is the dominant vector and *Plasmodium falciparum* is responsible for approximately 90% of all malaria cases (Máquina et al. 2024; Kloke et al. 2011; Aranda et al. 2005). Malaria transmission is highly seasonal, and varies across different regions in Mozambique, with the majority of cases occurring during the rainy season from November to April (Bertozzi-Villa et al. 2021; Ferrao et al. 2018). The analyses presented here thus provide insights relevant both for national malaria program planning and for broader climate–health modelling efforts.

Monthly counts of laboratory-confirmed malaria cases were obtained from the National Malaria Control Program (NMCP) and described in Harp et al. (2021). This data, reported through the *Boletín Epidemiológico Semanal*, covers the period from January 2017 to August 2023 and include all health facilities in 148 administrative districts. The fine spatial resolution of these aggregated data aligns well with the resolution of most global gridded climate observations. Per capita incidence values were computed using population estimates provided by the NMCP.

For this analysis, we focus on the ten districts with the highest mean malaria incidence during the study period (January 2017–August 2023). The geographic distribution of these districts is shown in Figure 4.

**Figure 4:**
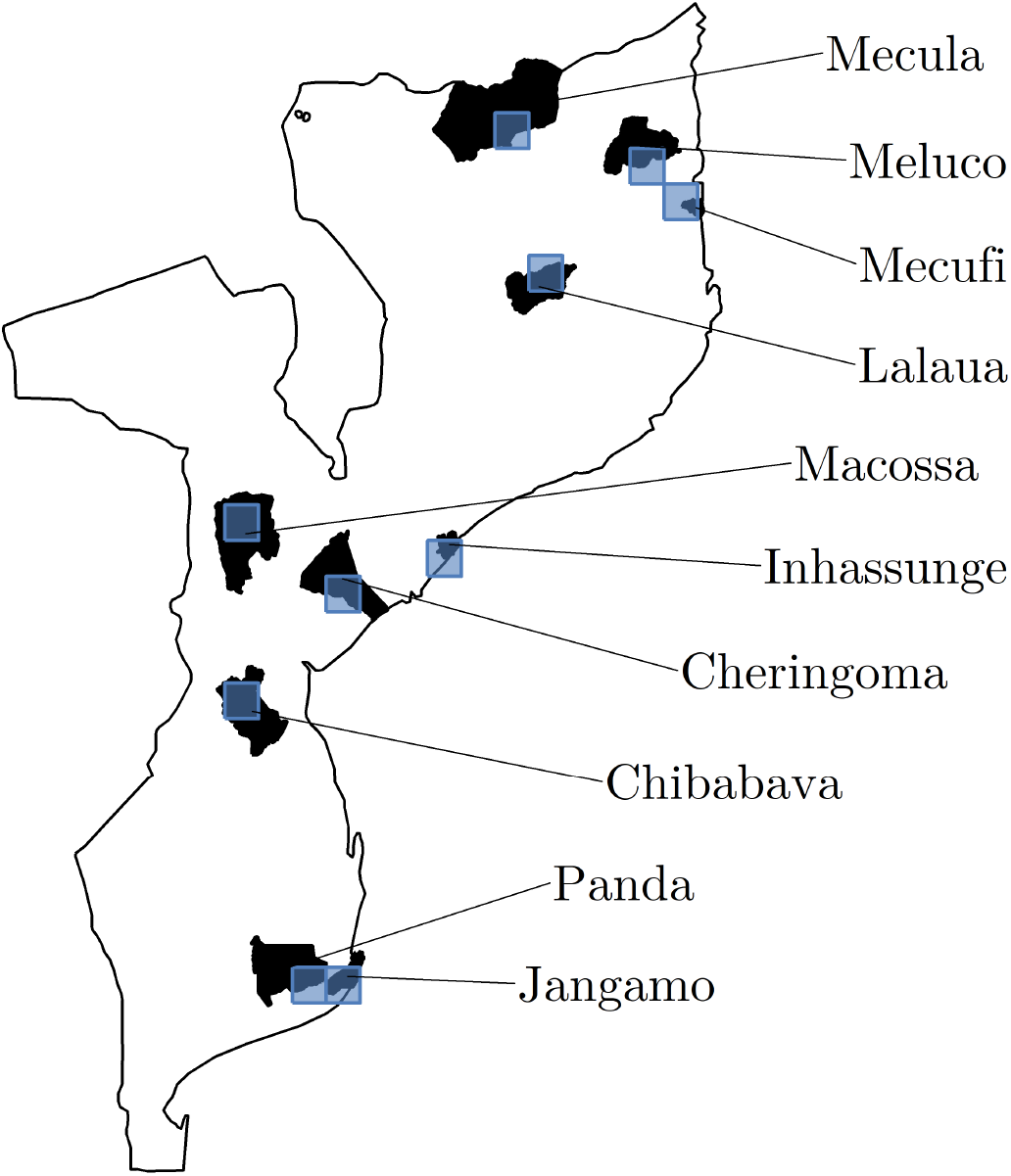
Map of the 10 districts in Mozambique with the highest incidence over the study period January 2017–August 2023. In black are the districts, and in blue the VECTRI grid cells representing these districts for which we extracted the climatic factors and run the VECTRI model. We selected the grid cell corresponding to each district by identifying the cell containing the district centroid.

We ran simulations using three model configurations: (1) The joint OpenMalaria-VECTRI modelling framework, as described in the previous section; (2) VECTRI-only, driven by the same climate and population density inputs as in the joint modelling framework; and (3) OpenMalaria-only, using an average EIR equal to 10% of the EIR produced by VECTRI.

For the OpenMalaria-only setup, we imposed a seasonal pattern of transmission based on monthly average rainfall, following OpenMalaria’s recommended approach. To account for the typical lag between rainfall and vector abundance, the rainfall-derived seasonality was shifted by one month (OpenMalaria 2026). For the OpenMalaria-only and the the joint OpenMalaria-VECTRI modelling framework, we replicated every simulation 100 times. For every output, we suppressed the outliers by removing the 5 highest and 5 lowest values, and calculated the mean of the remaining values.

Since the models produce outputs with different spatial and temporal units, we first harmonized the data before comparison. All simulated outputs were aggregated to a monthly resolution to match the temporal definition of the empirical data. We then rescaled model outputs to facilitate comparison, focusing on the timing, duration, and annual intensity of malaria transmission seasons rather than absolute values.

We extracted three main variables from the model outputs:

### a) Entomological Inoculation Rate (EIR)

For OpenMalaria and the joint modelling framework, we used the variable *simulatedEIR*. For VECTRI-only simulations, we used *eir*. To ensure comparability, the VECTRI EIR was rescaled to match the mean EIR of the joint modelling framework over the study period:

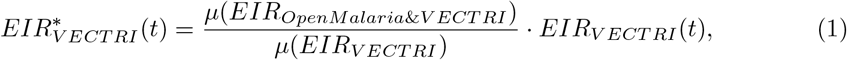

where *µ*(*·*) denotes the temporal mean over the simulation period.

### b) Malaria incidence

Empirical incidence in cases per 10,000 people per month was obtained from the NMCP data. For OpenMalaria and the joint modelling framework, we used the variable *nUncomp* (number of uncomplicated malaria episodes) as a proxy. For VECTRI, we used variable *cases*. Each malaria model’s simulated incidence was rescaled so that its mean matched the mean empirical incidence over the study period, similarly to the EIR calculation described in equation 1.

### c) Malaria prevalence

For OpenMalaria and the joint modelling framework, prevalence was derived from the variable *nPatent* (number of patent infections), that we divided by 10,000 people simulated (and later expressed in %). For VECTRI, we extracted the variable *PRd*. To ensure consistency, we scaled VECTRI to have the same prevalence mean as in the joint modelling framework, similarly as shown in equation 1.

To quantitatively assess the performance of the aforementioned models, we applied several complementary statistical analyses. To evaluate the timing of malaria incidence peaks, we employed a wavelet-based phase analysis following a method from Martin-Makowka et al. (2025). Specifically, we computed the cross-wavelet transform between the modeled and empirical incidence time series and extracted the phase difference at the dominant periodicity (that is, the 12-month period). The phase difference corresponds to the peak-to-peak time offset between the two time series, and can be expressed in months. When the modeled and empirical series were positively correlated, this phase difference directly represented the lag. When negatively correlated, half of the period (i.e., 6 months) was added to obtain the effective lag.

To assess the models ability to capture the interannual variability in malaria incidence, we compared modeled and empirical annual values for each district. For each model configuration, we plotted the empirical annual incidence against the modeled annual incidence and fitted a simple linear regression. Model accuracy is evaluated by observing whether the 1:1 line lays within the regression’s confidence interval. In addition, we computed the mean absolute error between empirical and modeled annual incidence for each district and model, providing a quantitative measure of deviation to the observed empirical value for annual incidence.

We also evaluated the ability of the models to reproduce the duration of the malaria transmission season. We defined this duration as the number of months per year with incidence values greater than or equal to the mean annual incidence. For each model and district, we plotted empirical versus modeled durations and applied a linear regression. Furthermore, we computed the coefficient of variation given by the standard deviation of monthly incidence normalized by its mean, for each district and modeled/empirical time series. This coefficient of variation served as a proxy for a seasonality index, quantifying the relative strength of variation in malaria incidence.

### Climate and effectiveness of indoor residual spraying

We examined how climate influences the timing and effectiveness of seasonal control interventions. The aim of this analysis was to assess whether interannual climatic variations can substantially alter the predicted timing and impact of seasonal interventions. Here, we specifically focus on the interplay between climate and indoor residual spraying (IRS).

For this modelling experiment, we selected the district of Chibabava in Mozambique, identified as the district with the highest malaria incidence (9,810 cases per 10,000 people per year) during the period January 2017–August 2023. Local climate data was extracted for this district, and simulations were conducted assuming an annual EIR of approximately 28 infectious bites per person per year (10% of the EIR suggested by VECTRI).

We first ran a standard OpenMalaria-only simulation in which the EIR seasonality was imposed following the rainfall seasonal pattern, shifted by one month. We iteratively repeated the simulations, deploying an IRS intervention once a year at time *t* (see below). The same set of simulations was then performed using the joint OpenMalaria-VECTRI modelling framework, with and without IRS deployment. For each simulation, we extracted the time series of simulated EIR (*simulatedEIR*), the number of uncomplicated malaria episodes (*nUncomp*), and the simulated malaria prevalence (*nPatent*).

When using identical input EIRs, minor discrepancies may arise between the simulated EIRs produced by the OpenMalaria-only and the joint OpenMalaria-VECTRI modelling framework due to nonlinear effects driven by interannual climate variations. To minimize such inconsistencies, we adjusted the input EIR for the joint modelling framework through an iterative, trial-and-error procedure to ensure comparable baseline transmission levels. In the absence of IRS, the adapted simulations produced mean EIRs of 28.087 and 27.559 infectious bites per person per year for the OpenMalaria-only and the joint OpenMalaria-VECTRI modelling framework, respectively.

IRS interventions were deployed annually at fixed times *t*, corresponding to either the 1st or 16th day of a given month. To avoid bias from the initial high-transmission period in early 2017, which would disproportionately benefit earlier deployment schedules, we started the first deployment time of IRS application after 6 months. The first IRS application was hence scheduled between July 1, 2017 and June 16, 2018. IRS intervention parameters followed the *IRS05* configuration described in Golumbeanu et al. (2024), which represents bendiocarb as the active ingredient tested against *Anopheles gambiae* (Chitnis et al. 2010). The decay in insecticidal concentration was modeled using an exponential function with a characteristic lifetime of *L* = 0.17 year and an initial postprandial killing effect of 0.81. The preprandial killing effect and deterrency were set to zero, and an IRS coverage of 90% was assumed. The intervention was deployed annually at the prescribed start times *t*.

The protective effect of IRS applied annually at time *t* was quantified relative to a no-intervention baseline. Protective factors were computed over the period from July 1st, 2017 to June 30th, 2023, for both transmission and disease outcomes: (i) the number of infectious bites averted, and (ii) the number of uncomplicated malaria episodes averted. The protective factor against malaria incidence was defined as:

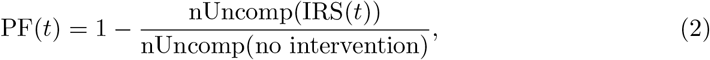

where, nUncomp(IRS(*t*)) represents the total number of uncomplicated malaria episodes in the simulation with IRS deployed at time *t*, and nUncomp(no intervention) represents the corresponding number without IRS. A similar computation was applied to EIR to estimate the protective factor against disease transmission.

## Results

### Empirical malaria incidence

We first examined the temporal evolution of empirical malaria incidence data from Mozambique. The spatial and temporal distribution of incidence for the ten districts with the highest malaria burden during the study period (January 2017–August 2023) is shown in Figure 5. These districts were selected because they collectively account for the majority of reported malaria cases nationwide.

**Figure 5:**
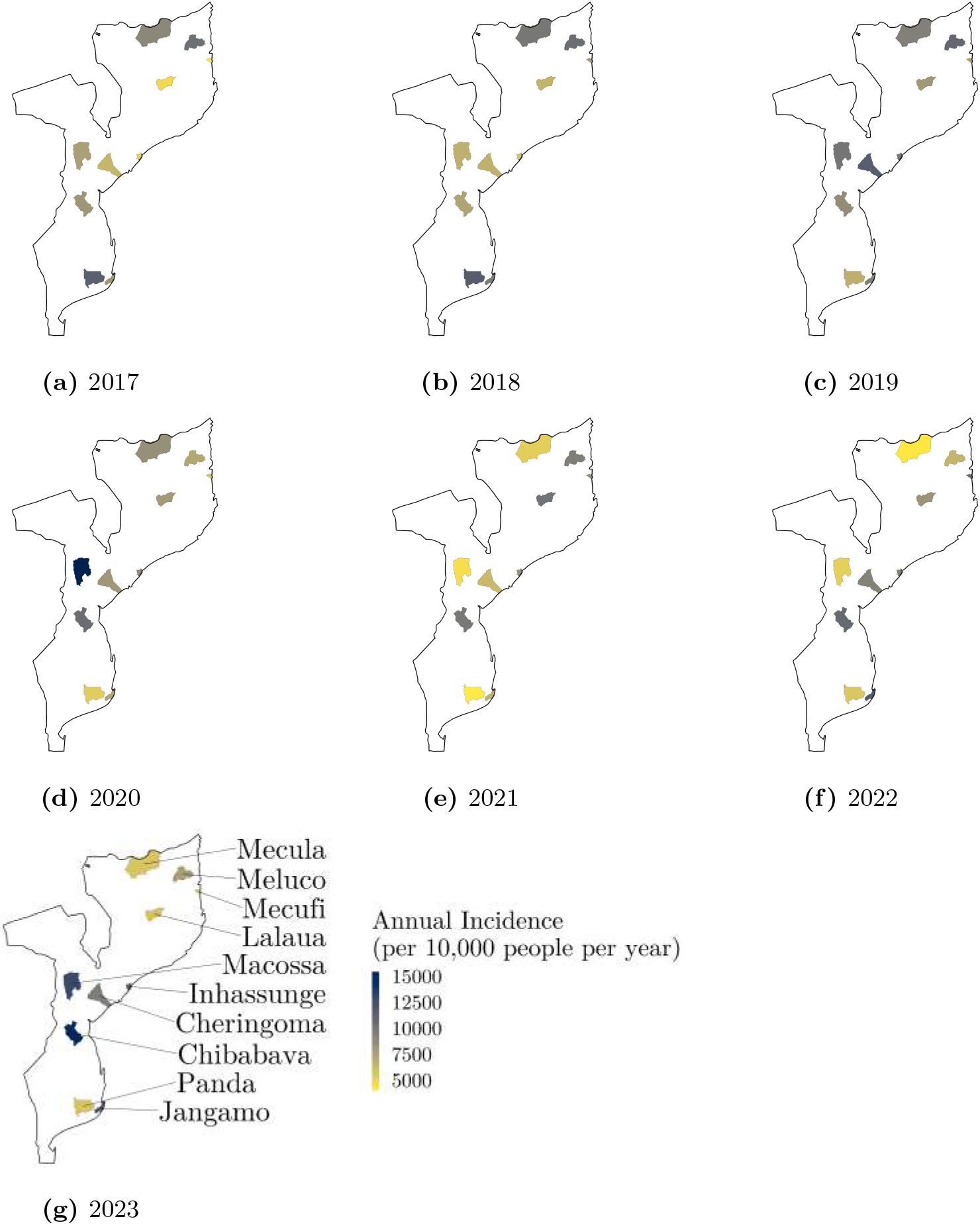
Evolution of malaria incidence from 2017 to 2023, in the 10 districts bearing the highest incidence burden. The incidence is computed as the total number of confirmed malaria cases per 10,000 people and per year. The data for 2023 only ranges between January and August, and the annual incidence has been scaled by a corresponding factor.

The analysis reveals substantial spatial heterogeneity in malaria trends across Mozambique. Several districts located in the northern and southern regions (Mecula, Meluco, and Panda) exhibit a clear decreasing trend in annual malaria incidence over time. In contrast, smaller coastal districts such as Mecufi, Jangamo, and Inhassunge show no consistent longterm trend but share a similar temporal pattern, with distinct incidence peaks observed in 2019, 2022, and 2023. The district of Lalaua, located in the central-northeastern region, shows an increase in malaria incidence around 2021 followed by a gradual decline toward the end of the study period. The two central-northern districts, Macossa and Cheringoma, both display bimodal patterns, with elevated incidence levels in 2019–2020 and again in 2023. The district of Chibabava in the center-south stands out as the area with the highest malaria incidence overall. It exhibits an upward trend in malaria incidence throughout the study period.

### Model validation

We validated the performance of the OpenMalaria-only and VECTRI-only simulations, together with the joint OpenMalaria-VECTRI modelling framework. The modeled time series for the Chibabava district are presented in Figure 6, while results for the remaining districts are provided in the Appendix (Figures S1–S3).

**Figure 6:**
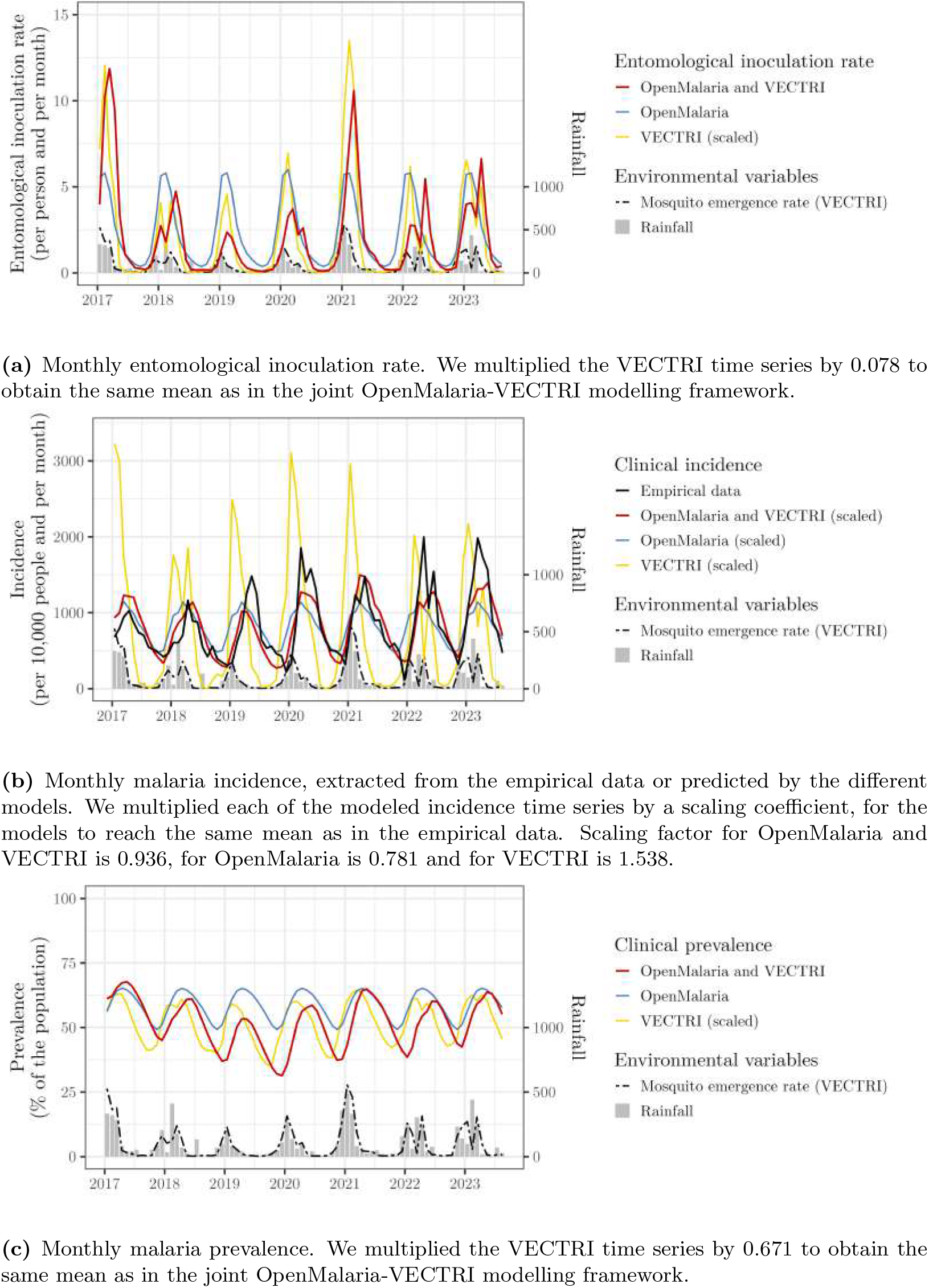
Comparison between modeled or empirical malaria metrics for the district of Chibabava. Times series represent the a) entomological inoculation rate, b) incidence, c) prevalence, as well as monthly rainfall (mm) and VECTRI-computed emergence rate.

Figure 6a shows the evolution of the EIR across models, together with associated rainfall and VECTRI-simulated mosquito emergence rate, allowing us to relate transmission dynamics to climatic forcing. Rainfall in Chibabava is generally unimodal, with an austral summer rainy season lasting from December to March and peaking around January, although bimodal patterns are also observed in 2018, 2022, and 2023. Occasionally, isolated rainfall peaks can occur, such as in July 2018. Simulated mosquito emergence rate closely follows rainfall seasonal cycles, typically without lag, although in 2018, 2022, and 2023, a one-month lag is observed. In VECTRI-only simulations, the EIR peaks are delayed by one month with respect to the emergence rate peaks. In OpenMalaria-only simulations, EIR patterns are highly regular: for years with a single rainfall peak, EIR closely aligns with the VECTRI peak, whereas for years with multiple rainfall peaks (2018, 2022, 2023), a single EIR peak occurs, approximately representing an average of the rainfall peaks predicted by VECTRI. In the joint OpenMalaria-VECTRI modelling framework, EIR peaks tend to coincide either with the VECTRI-only timing or occasionally occur one month later.

The modeled and empirical malaria incidence time series for Chibabava are compared in Figure 6b. The empirical data show an overall intensification of malaria transmission toward the end of the study period, with predominantly unimodal seasonal cycles peaking between March and May each year. Exceptions include a bimodal season in 2022, a moderate rebound during 2020–2021, and a small secondary increase in September 2018. The lag between VECTRI-simulated emergence and empirical incidence varies between 1 and 4 months, typically around 2 months. In general, VECTRI tends to lead the empirical signal by approximately two months, with simulated cases peaking concurrently with emergence rate, except in 2018, 2022, and 2023, where a one-month lag occurs. The OpenMalaria-only simulations show regular seasonal patterns with annual peaks around March, slightly ahead of the empirical data. These simulations reproduce the seasonal cycles but with weaker contrasts: the increase in incidence at the start of the season is more gradual, and the difference between high and low seasons is attenuated. The lag between emergence and incidence is again around two months. The joint OpenMalaria-VECTRI modelling framework captures the onset of the transmission season well and follows the empirical timing more closely than standalone models, although it tends to lead slightly in 2018–2019. As for the other simulations, no clear long-term trend is observed. In such configuration, incidence typically peaks about two months after the simulated emergence rate peak.

Prevalence time series for Chibabava are shown in Figure 6c. No empirical prevalence data is available for direct comparison, and the analysis focuses on differences between models. VECTRI consistently predicts earlier peaks in prevalence than the other configurations, typically preceding the OpenMalaria-only peaks by about one month and the joint OpenMalaria-VECTRI modelling framework by two months. Modeled prevalence fluctuates seasonally between roughly 40% and nearly 100%, depending on the year and model setup.

We further assessed the performance of the OpenMalaria-only, VECTRI-only, and joint OpenMalaria-VECTRI modelling framework using a suite of statistical analyses. Results for the Chibabava district are illustrated in Figure 7, while the corresponding analyses for the remaining districts are shown in the Appendix (Figures S4, S5, and S6).

**Figure 7:**
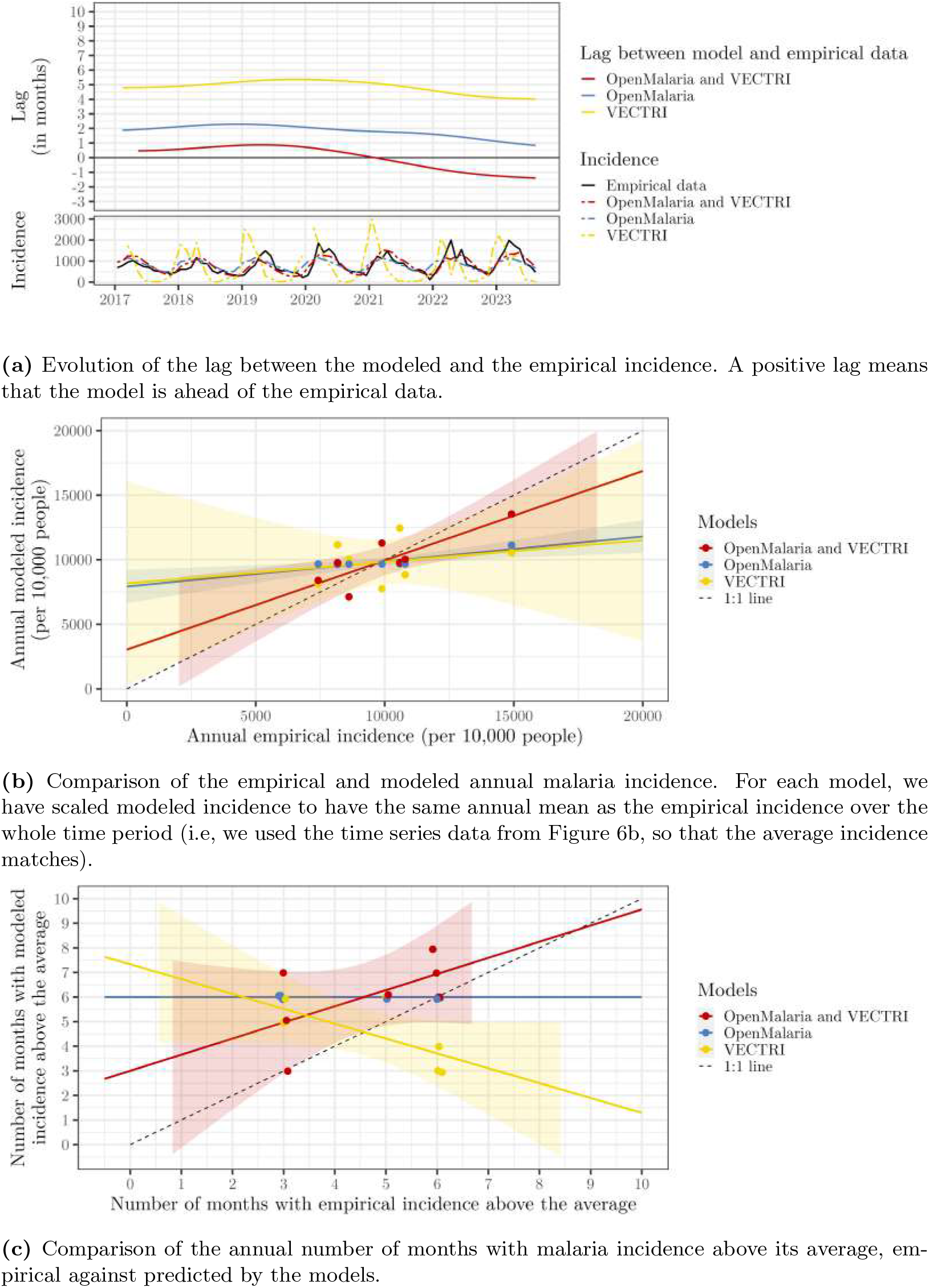
Evaluation of the performance of the 3 different models. (VECTRI, OpenMalaria and OpenMalaria-VECTRI) in the district of Chibabava.

We first quantified the timing accuracy of each model by estimating the lag between the modeled and empirical incidence time series using a wavelet-based phase-difference approach. This method captures the temporal evolution of lags across the study period and focuses on the dominant 12-month periodicity. Results for Chibabava are presented in Table 7a. For VECTRI, the modeled incidence leads the empirical signal by approximately five months at the beginning of the study period, decreasing monotonically to about four months by the end. The OpenMalaria-only model initially leads the empirical data by roughly two months, with the lag gradually reducing to slightly less than one month toward the end of the period. In contrast, the joint OpenMalaria-VECTRI modelling framework shows much smaller lags: it begins with a positive lag of about half a month (model slightly ahead), crosses zero (in synchrony) in early 2021, and ends with a negative lag of approximately−1 month, indicating a minor delay relative to observations.

We then compared the models’ ability to reproduce annual incidence totals using scatter plots and linear regressions in Figure 7b. The VECTRI-only model shows large interannual variability, resulting in a wide confidence interval that does not include the 1:1 line. The OpenMalaria-only model exhibits no interannual variation and predicts nearly identical incidence every year, yielding a very narrow and flat regression confidence interval with a near-zero slope. In contrast, the joint OpenMalaria-VECTRI modelling framework produces the steepest regression slope, and its confidence interval includes the 1:1 line, indicating that it better captures interannual changes in malaria incidence.

We also evaluated the ability of each model to reproduce the duration of the malaria transmission season. Duration was defined as the number of months per year with incidence greater than or equal to the annual mean (Figure 7c). For VECTRI, the regression between modeled and empirical duration has a negative slope, indicating that the model predicts shorter seasons in years with higher empirical incidence. The OpenMalaria-only model again produces nearly constant estimates each year, resulting in a flat slope and no meaningful confidence interval. The joint OpenMalaria-VECTRI modelling framework displays substantial interannual dispersion, with simulated transmission seasons ranging from 3 to 8 months; however, its confidence interval includes almost the entire 1:1 line, indicating satisfactory agreement with empirical values.

Table 1 shows the mean lag (and its standard deviation) between modeled and empirical incidence for each district. For both VECTRI-only and OpenMalaria-only, all lags remain strictly positive, reflecting consistent anticipation of the empirical signal across districts. For each district, the model with lag closest to zero is highlighted in bold. Table 2 presents the mean absolute error (MAE) between observed annual incidence and the corresponding simulated annual values. The smallest MAE for each district is highlighted in bold. Finally, Table 3 shows the standard deviation of the monthly time series—used here as a seasonality index reflecting how strongly incidence fluctuates within each year. For each district, the simulated value closest to the empirical standard deviation is highlighted in bold.

**Table 1:** Mean lags across models and districts, computed from the phase differences.

| district | VECTRI | OpenMalaria | OpenMalaria and VECTRI |
| --- | --- | --- | --- |
| Chibabava | 4.85 | 1.81 | **0.01** |
| Jangamo | 4.01 | **0.57** | -0.91 |
| Meluco | 6.65 | 4.74 | **2.44** |
| Macossa | 3.12 | 1.67 | **-0.92** |
| Inhassunge | 8.74 | 3.96 | **2.43** |
| Cheringoma | 4.51 | 1.75 | **-0.31** |
| Panda | 3.16 | 2.71 | **1.24** |
| Lalaua | 3.28 | **0.61** | -1.16 |
| Mecula | 2.41 | 1.52 | **-1.33** |
| Mecufi | 3.11 | **0.14** | -1.84 |

**Table 2:** Mean absolute errors between the predicted and observed annual incidence, across models and districts.

| district | VECTRI | OpenMalaria | OpenMalaria and VECTRI |
| --- | --- | --- | --- |
| Chibabava | 2205.61 | 1539.08 | **1201.20** |
| Jangamo | 2698.03 | 1540.85 | **1302.14** |
| Meluco | 1919.21 | **1871.68** | 1989.23 |
| Macossa | **2891.25** | 2970.39 | 3589.54 |
| Inhassunge | 2242.16 | 1704.17 | **1548.63** |
| Cheringoma | 1824.82 | **1384.82** | 1561.90 |
| Panda | **1386.03** | 2557.51 | 2825.62 |
| Lalaua | 2238.58 | 1702.31 | **1488.39** |
| Mecula | **1995.09** | 2351.82 | 2751.85 |
| Mecufi | 1736.88 | 1894.02 | **1578.13** |

**Table 3:** Coefficients of variation of the modeled and empirical incidence time series normalized by mean, per district and model. Standard deviation is used as a proxy for seasonality index including the interannual variation (how well malaria transmissions seasons are marked).

| district | VECTRI | OpenMalaria | OpenMalaria and VECTRI | Empirical |
| --- | --- | --- | --- | --- |
| Chibabava | 1.08 | 0.27 | **0.43** | 0.53 |
| Jangamo | 0.85 | 0.19 | **0.23** | 0.34 |
| Meluco | 0.83 | **0.39** | 0.35 | 0.39 |
| Macossa | 1.23 | 0.41 | **0.68** | 0.76 |
| Inhassunge | 0.98 | 0.21 | **0.27** | 0.35 |
| Cheringoma | 1.06 | 0.25 | **0.43** | 0.52 |
| Panda | 0.92 | 0.18 | **0.28** | 0.52 |
| Lalaua | 0.87 | 0.48 | **0.39** | 0.38 |
| Mecula | 1.21 | **0.51** | 0.66 | 0.43 |
| Mecufi | 0.82 | **0.37** | 0.31 | 0.52 |

### Impact of climate on the effectiveness of indoor residual spraying

Following validation of the joint OpenMalaria-VECTRI modelling framework, we now explore the simulated interplay between climate and malaria intervention dynamics. Figure 8 presents the results for a scenario where IRS is deployed annually on November 16. The intervention produces a marked reduction in malaria transmission, reflected by a strong decrease in simulated EIR and prevalence. The reduction in malaria incidence is less pro-nounced but still noticeable. As expected, the concentration of active ingredients in the IRS intervention is highest immediately following deployment and declines rapidly thereafter, consistent with the exponential decay defined by the model parameterization.

**Figure 8:**
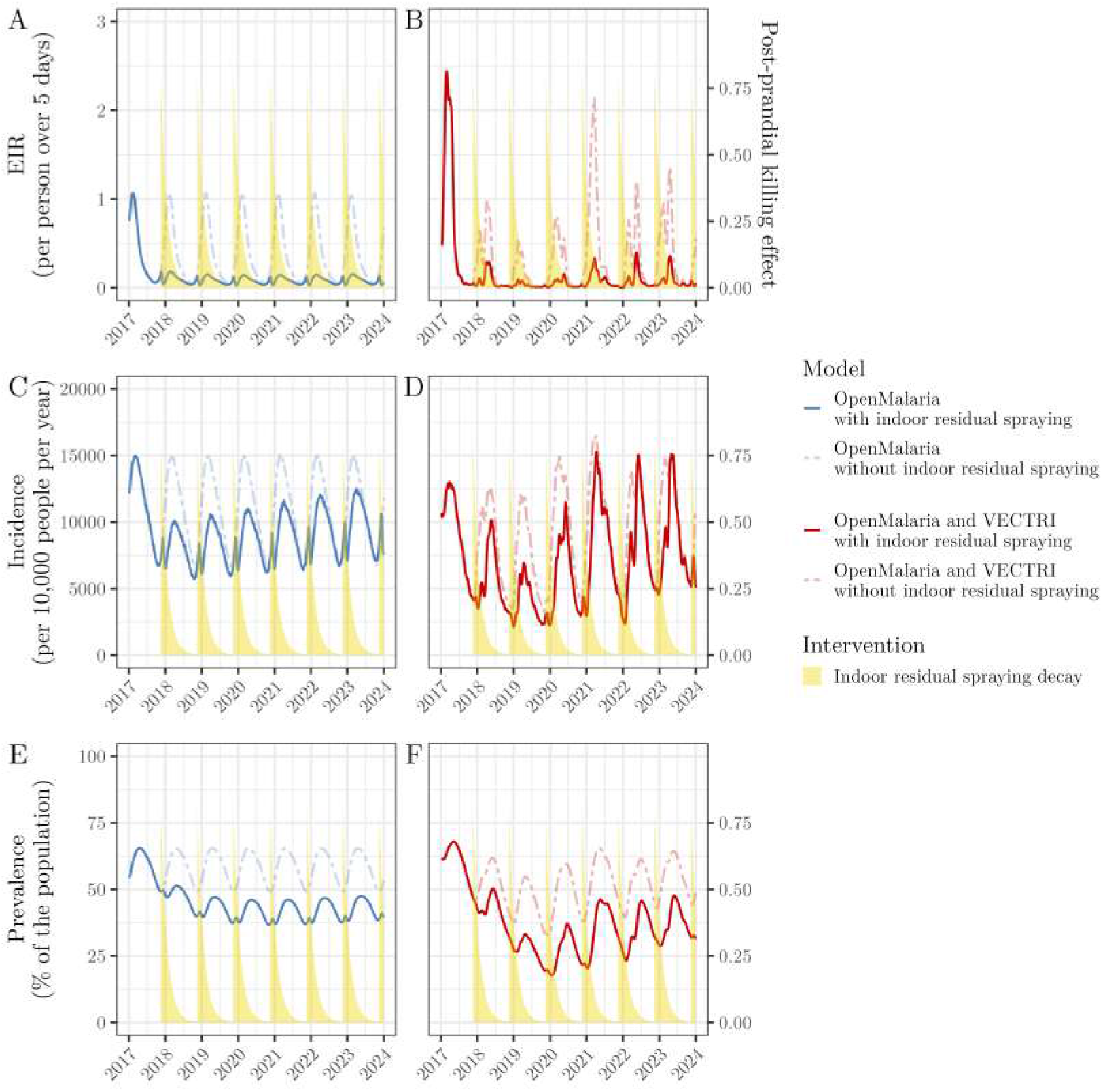
Simulated effect of the deployment of indoor residual spraying on different malaria metrics, using the standard version of OpenMalaria or the joint OpenMalaria-VECTRI modelling framework. Indoor residual spraying is deployed every year on November 16. Study duration spans from January 2017 to December 2023. Plots A and B show the predicted effect of both models on the entomological inoculation rate (new infectious bites per person over the previous 5 days), plots C and D on the incidence (new episodes in the previous 5 days, expressed as of per year per 10,000 people), and plots E and F on the prevalence (% of the total population). Areas in yellow represent the deployment and decay of the post-prandial killing effect of the indoor residual spraying. On this plot, none of the metrics have been rescaled, hence the output cannot be compared to Figure 6.

In the OpenMalaria-only simulations, transmission patterns remain identical from year to year, since interannual variability is not explicitly represented. In contrast, the joint OpenMalaria-VECTRI modelling framework incorporates interannual fluctuations in climate, resulting in year-to-year variability in simulated EIR, incidence, and prevalence, even under identical intervention schedules.

Figure 9a summarizes the protective factor of IRS against EIR as a function of deployment timing and model. In the OpenMalaria-only simulations, IRS achieves an average protective factor of approximately 77.62%, corresponding to a 22.38% residual transmission relative to the non-intervention scenario. The optimal deployment timing in this case is December 1. In comparison, the joint OpenMalaria-VECTRI modelling framework predicts a stronger protective factor of 82.30%, reducing transmission to 17.70% of its original value. The maximum effectiveness is achieved when IRS is deployed on December 16 each year.

**Figure 9:**
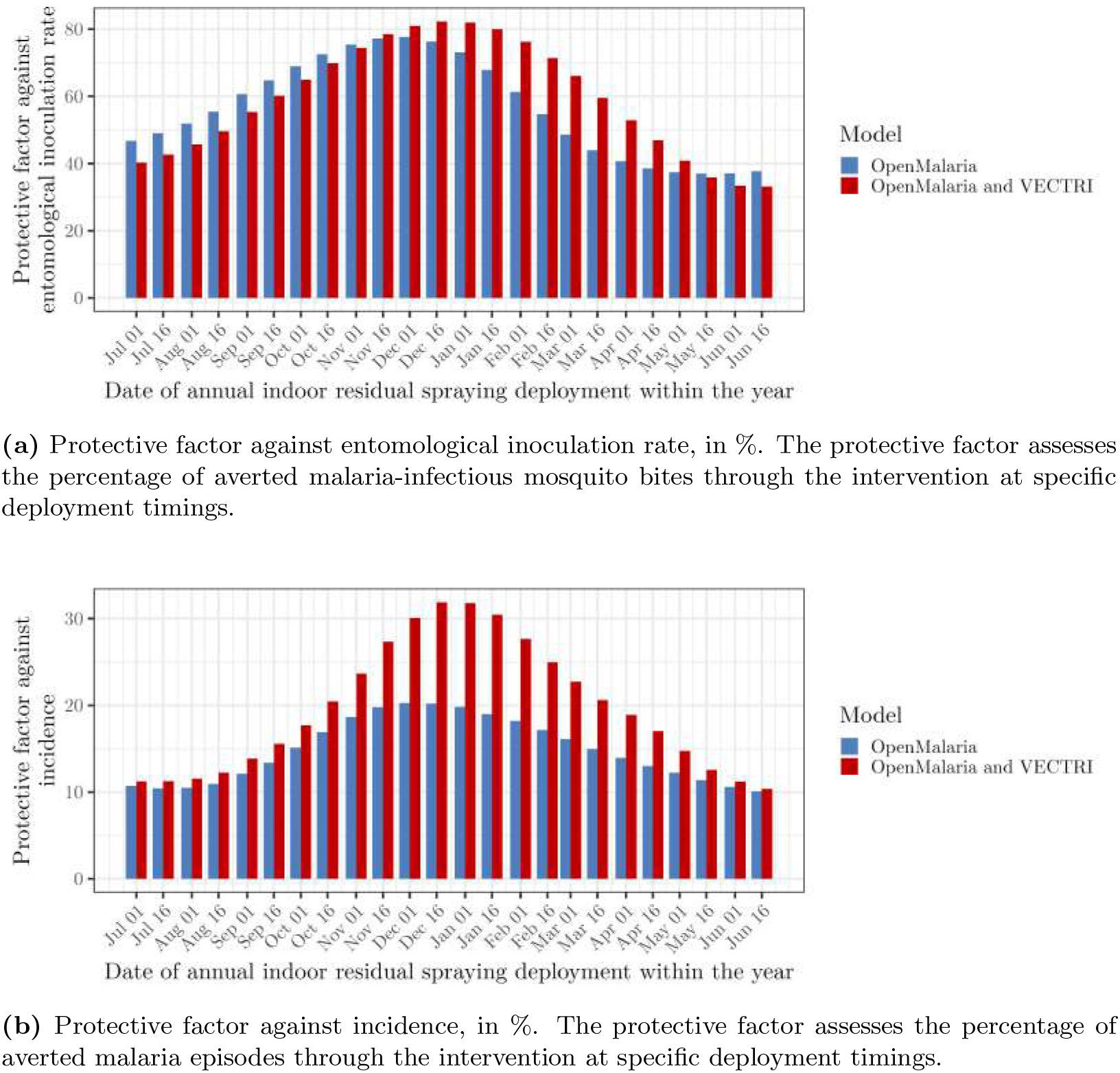
Protective factors of the indoor residual spraying interventions, depending on the selected model (standard OpenMalaria or the joint OpenMalaria-VECTRI modelling framework) and the date of the year when the indoor residual spraying intervention is deployed (Figure 8 is an example for indoor residual spraying deployment on November 16).

The protective factor against malaria incidence, representing the proportion of malaria episodes averted through IRS deployment, is shown in Figure 9b. The OpenMalaria-only model estimates an optimal deployment on December 1, yielding a protective factor of 20.28%. In contrast, the joint OpenMalaria-VECTRI modelling framework simulations indicate that the most favorable deployment timing occurs around December 16, with a protective factor of 31.88%.

## Discussion

In this study, we developed and evaluated a new malaria modelling framework that couples the climate-driven model VECTRI with the individual-based epidemiological model OpenMalaria. Our aim was to construct a modelling workflow capable of capturing interannual, climate-driven variability in malaria transmission while retaining the detailed representation of within-host dynamics and intervention effects implemented in OpenMalaria. We validated this framework using malaria incidence data from the highest-incidence districts in Mozambique. In addition, we assessed how interannual climatic variability influences the predicted effectiveness of seasonally-deployed IRS.

Validation shows that the joint OpenMalaria-VECTRI modelling framework accurately captures the timing of malaria incidence peaks. In the district of Chibabava, the lag between the empirical incidence time series and the OpenMalaria-VECTRI incidence time series never exceeds one month, well below the OpenMalaria-only and the VECTRI-only models. When extended to all districts, the joint modelling framework performs best in 7 out of 10 high-incidence districts, achieving average lags closest to zero. The OpenMalaria-only model performs slightly better in the remaining districts, suggesting that there is no universal one-size-fits-all model. Nonetheless, the overall improvement is substantial; while single models sometimes exhibit large timing errors, the joint modelling framework keeps average empirical–model incidence lags below 2.5 months. In the districts where the models perform poorly, the districts with the largest remaining discrepancies (Meluco, Inhassunge, and Mecúfi) share either weak seasonal rainfall regimes (Inhassunge) or strong rainfall seasonality but no consistent malaria seasonality (Meluco, Mecúfi). All three are coastal districts that may experience strong, irregular climatic shocks, including cyclones, flooding and surges. These events can influence mosquito ecology or health-facility reporting in ways that are not captured by rainfall and temperature data alone. Conversely, districts with clear rainfall seasonality, such as Chibabava, Macossa, Cheringoma, and Mecula, show highly accurate timing predictions under the joint modelling framework.

Observed lags between rainfall and malaria incidence in Mozambique and other African settings typically range from one to two months (Beloconi et al. 2023; Nyawanda et al. 2023; Ferrão et al. 2017), depending on rainfall intensity (Armando et al. 2023). A two-month lead time is generally considered optimal for early-warning systems (Armando et al. 2025). This reflects the cumulative time required for mosquito breeding, sporogony, and subsequent human infection (Reiner et al. 2015). Our joint modelling approach reproduces this characteristic lag structure, the occurrence of rainfall leading the malaria incidence peak by approximately two months.

Model performance on annual incidence values is less clear-cut. While the joint OpenMalaria-VECTRI modelling framework performs well in Chibabava and outperforms single models in 5 out of 10 districts (notably in Macossa, Panda, and Mecula), prediction errors remain substantial (1,200–3,600 cases per 10,000 people). Both the joint and single models struggle to reproduce district-specific long-term trends, which may stem from temporal changes in intervention coverage, health-system functionality, socio-economic factors, or human mobility—factors not yet represented in the model.

The joint modelling framework successfully captures the duration of the transmission season in Chibabava, with positive correlations between empirical and simulated season lengths. Across districts, it also better reproduces the seasonality index (the standard deviation of monthly incidence) compared with single models, performing best in 7 out of 10 districts. These results align with the dominant role of rainfall and temperature in shaping malaria seasons (Reiner et al. 2015). With our novel modelling approach integrating climate-driven mosquito dynamics from VECTRI and accurate immunity profiles from OpenMalaria, the joint modelling framework effectively captured the strength of both models.

Overall, the joint modelling framework shows clear improvements over single models, particularly for predicting timing and season duration, while improvements in intensity prediction remain more modest.

Because the joint OpenMalaria-VECTRI modelling framework performs particularly well in highly seasonal, high-incidence settings such as Chibabava, it provides a meaningful basis for evaluating seasonal interventions such as IRS. In these simulations, IRS substantially reduced the EIR, consistent with its mode of action targeting indoor-resting, human-fed mosquitoes. The impact on incidence and prevalence is more modest. Reduction in EIR also reduces population-level immunity, which dampens the downstream reduction in clinical cases. Under optimal deployment timing, OpenMalaria-only predicts EIR reduction of 77.62% and incidence reduction of 20.28%, whereas the joint OpenMalaria-VECTRI modelling framework predicts larger effects of 82.30% and 31.88%, respectively. The joint modelling framework aligns closer to empirical findings that are typically show an effectiveness of annually-deployed IRS intervention campaigns on malaria incidence of around 30% (Hilton et al. 2023). The inclusion of climate variables enables the model to reproduce the near-absence of transmission during the dry season, with a sharper increase at the beginning of the transmission season. In contrast, standard OpenMalaria assumes an idealized seasonal forcing with persistent low-level transmission risk year-round, which underestimates the predicted benefit of interventions. Thus, accounting for climate not only improves simulations but also yields different and arguably more accurate estimates of optimal intervention timing and expected effectiveness.

The optimal IRS deployment timing should occur in early December according to OpenMalaria-only and mid-December for the joint OpenMalaria-VECTRI modelling framework. The malaria transmission timing being improved in the joint OpenMalaria-VECTRI modelling framework, we can argue that this difference reflects the improved representation of transmission season timing in the joint modelling approach.

Although our novel modelling approach fit malaria transmission seasons closer and predicts expected protective efficacy better, limitations remain. Namely, none of the models could successfully catch the trends in malaria incidence. While some variations of malaria incidence in Mozambique are influenced directly by rainfall and temperature, they may be also attributed to the effects of external factors. Mozambique deploys insecticide-treated nets in repeated mass distribution cycles, typically every three years, which can strongly influence interannual incidence patterns (Wetzler et al. 2022). At the same time, the country is highly exposed to extreme climatic events, including the impact of cyclones and floods. Mozambique was hit in 2017 by cyclone Dineo, in 2019 by cyclones Idai and Kenneth and in 2023 by cyclone Freddy (Rossi et al. 2024). These shocks are not always captured by monthly mean rainfall or temperature time series but can nevertheless disrupt malaria transmission or the reporting thereof. For instance, cyclone Freddy in 2023 caused extensive damages to health systems and may have altered malaria case detection, treatment availability, and human exposure patterns. Such climatic and non-climatic disturbances create complex incidence trends that can be challenging for mechanistic models to capture, especially when models are solely driven by rainfall and temperature data, as it is the case for our malaria modelling framework.

Mosquito emergence in the current joint modelling approach is not directly affected by vector control interventions. In OpenMalaria, emergence is seasonally prescribed and independent of adult mosquito density, whereas in VECTRI it depends on larval dynamics and adult populations that directly rely on rainfall and temperature inputs. Ideally, the models should be dynamically coupled: OpenMalaria should pass intervention-induced mosquito mortality back to VECTRI, allowing VECTRI to adjust emergence rates dynamically. Implementing such two-way coupling is a priority for future work.

The effective EIR used in the simulations was fixed at 10% of the value suggested by VECTRI. This choice was made to account for the different level of transmission intensity suggested by VECTRI. Taking only a fraction of the EIR was needed to obtain a more realistic transmission level, consistent with typical EIRs in Mozambique. Similarly, modeled incidence had to be rescaled to match empirical incidence. The primary objective of this piece of work was to evaluate whether the joint modelling framework could reproduce the timing and structure of transmission seasons rather than absolute incidence levels. Ideally, the modelling framework should allow direct calibration of transmission intensity, enabling EIR to be adjusted so that key epidemiological outputs, such as incidence and prevalence, match empirical targets. Developing such an internally consistent calibration strategy represents an important next step to strengthen the quantitative interpretability of the framework.

In conclusion, we have developed a novel modelling framework that merges climate-driven mosquito dynamics from VECTRI with the detailed human infection dynamics and intervention modelling of OpenMalaria. This is the first framework to combine mechanistic climate-driven transmission dynamics with detailed modelling of intervention effectiveness. This approach captures interannual climate variability and improves predictions of malaria transmission timing and seasonality in high-incidence districts of Mozambique. When applied to seasonal interventions, our modelling framework reveals significant differences in the optimal deployment timing and impact of IRS compared to the standard OpenMalaria model, underscoring the importance of representing climate and interannual variability in malaria control assessments. The joint OpenMalaria-VECTRI modelling framework offers a promising platform for future analyses and for designing more context-specific malaria intervention strategies.

## Acknowledgments

The authors thank Dr. Baltazar Candrinho, Director of the National Malaria Control Program of Mozambique, for the malaria incidence data used to validate the modelling framework. The authors also acknowledge Kevin Crowe, data analyst at The Washington Post, for facilitating access to these data.

A.M.M. and N.C. acknowledge support from the Gates Foundation [Grant number 025569 (A.M.M. and N.C.) and 081779 (N.C.)].

A.M.M., J.D.M., A.P.M., C.C., N.C. acknowledge support from the Wellcome Trust CLIMate SEnsitive DISease forecasting tool (CLIMSEDIS) research project [Grant number 225997/Z/22/Z].

## Appendix

**Figure S1:**
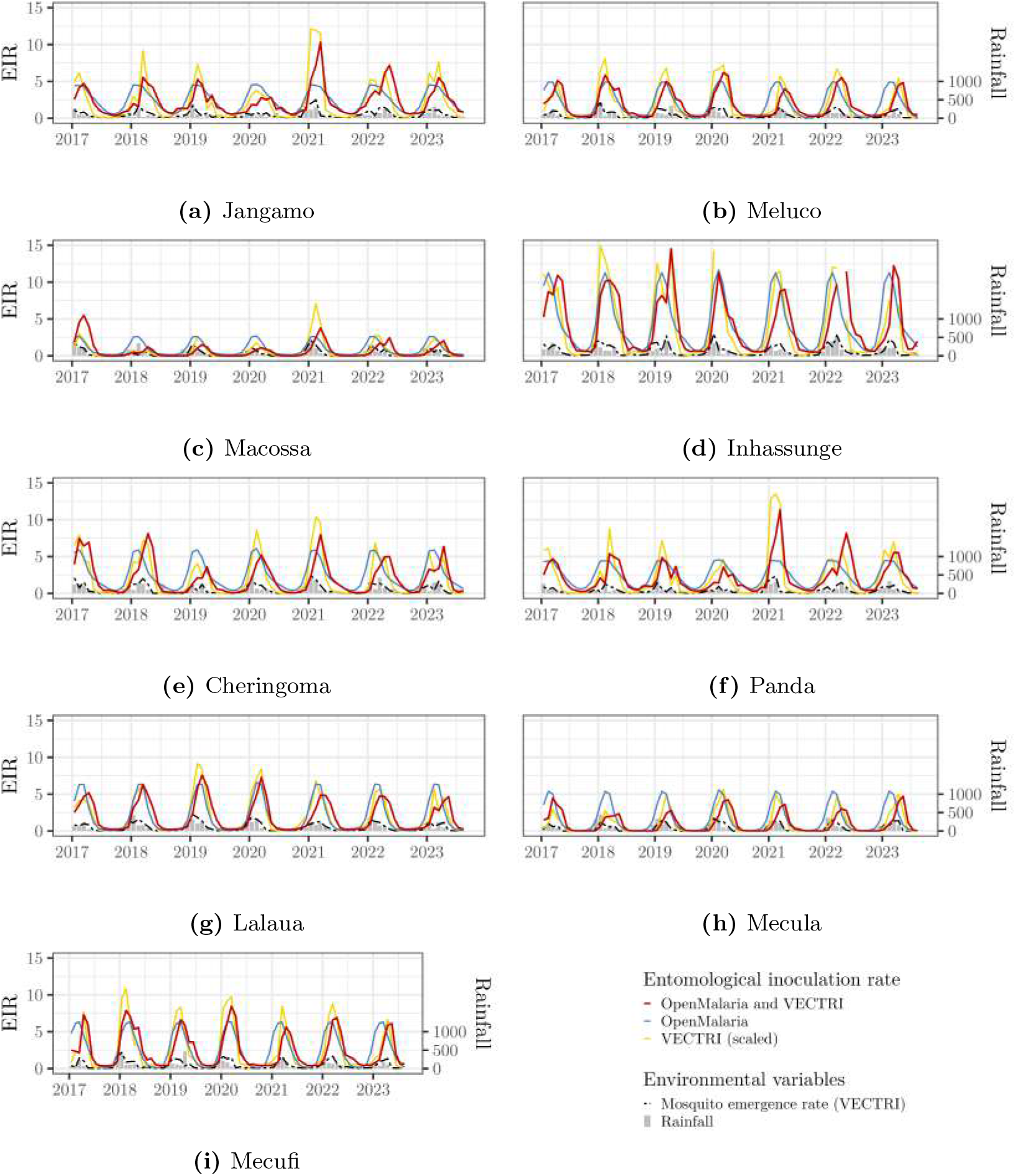
Monthly entomological inoculation rate, as predicted by the different models, for the 9 Mozambican districts with the highest incidence after Chibabava (shown in the main article). We transformed the variables ‘simulatedEIR’ (OpenMalaria, and OpenMalaria-VECTRI) and ‘eir’ (VECTRI) into a monthly entomological inoculation rate. We multiplied the monthly entomological inoculation rate from VECTRI by a scaling coefficient to obtain the same mean between the VECTRI and the OpenMalaria-VECTRI models. We plotted the monthly rainfall to display the rainy seasons. We plotted the VECTRI-modeled emergence rate to show the intermediate step in both the VECTRI and the joint OpenMalaria-VECTRI modelling framework.

**Figure S2:**
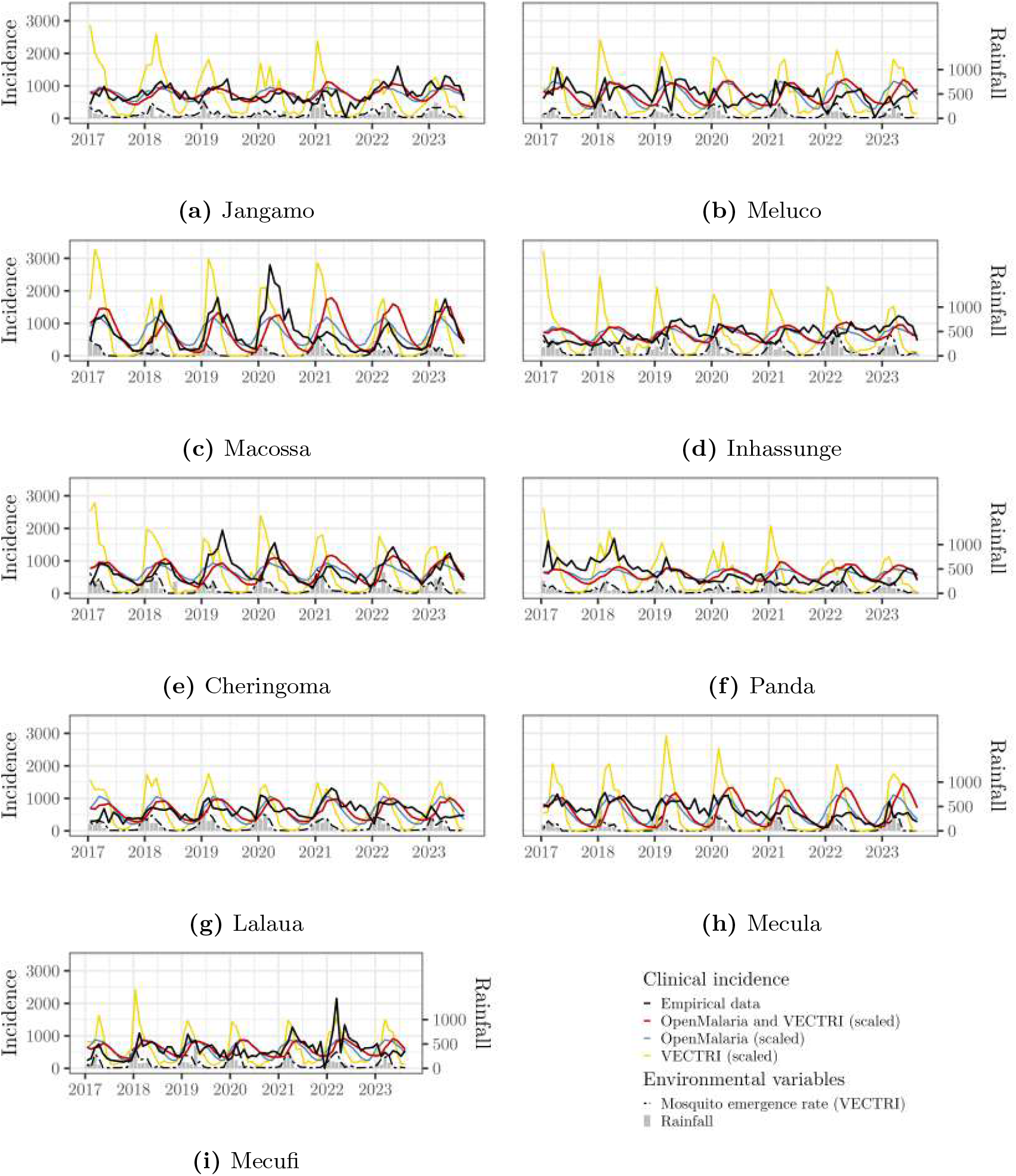
Monthly malaria incidence, extracted from the empirical data or predicted by the different models, for the 9 Mozambican districts with the highest incidence after Chibabava (shown in the main article). We transformed the variables ‘nUncomp’ (OpenMalaria, and OpenMalaria-VECTRI) and ‘cases’ (VECTRI) into a monthly incidence. We multiplied each of the modeled incidence time series by a scaling coefficient, to obtain the same mean between all models and the empirical data. We plotted the monthly rainfall to display the rainy seasons. We plotted the VECTRI-modeled emergence rate to show the intermediate step in both the VECTRI and the joint OpenMalaria-VECTRI modelling framework.

**Figure S3:**
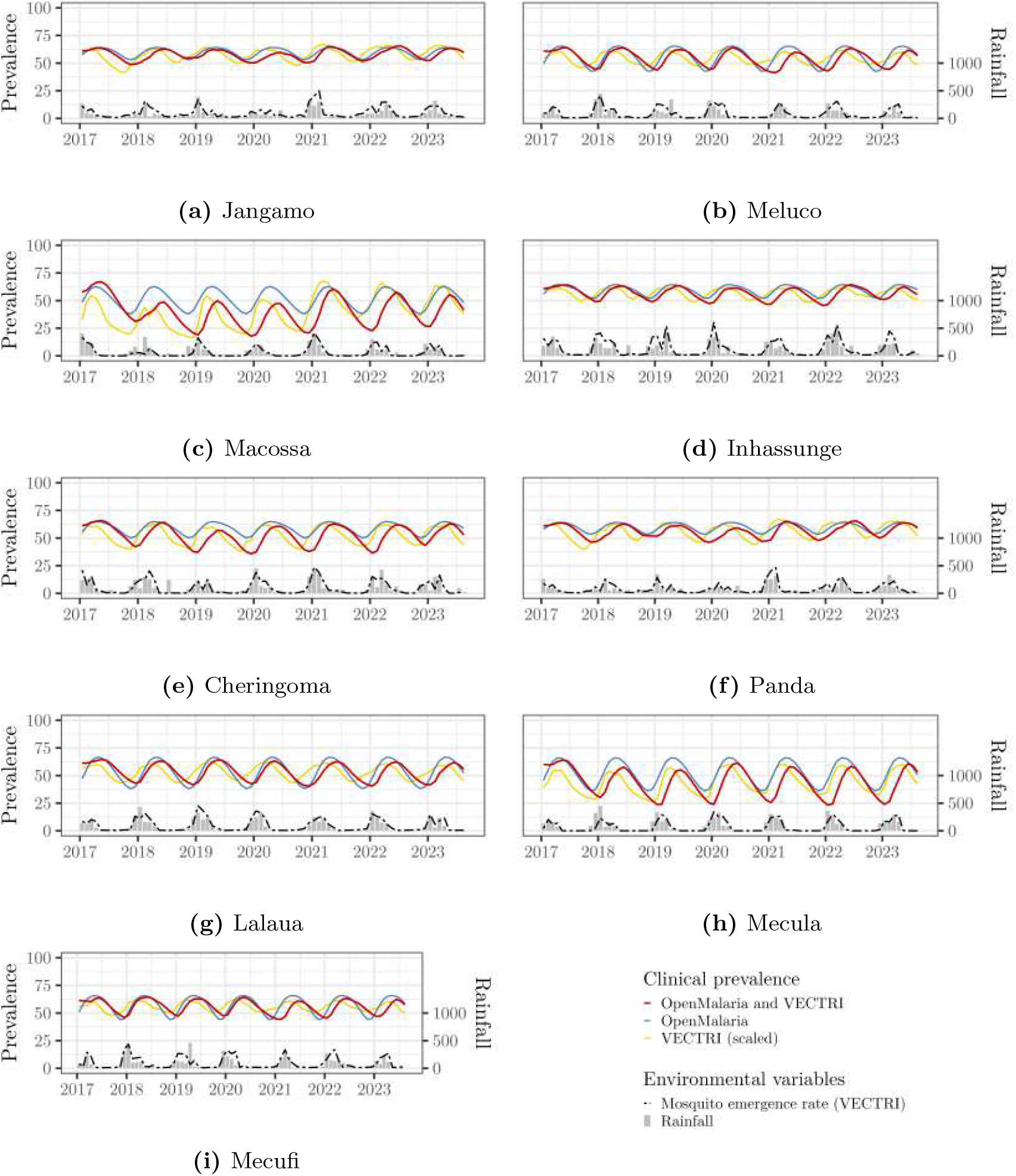
Monthly malaria prevalence, as predicted by the different models, for the 9 Mozambican districts with the highest incidence after Chibabava (shown in the main article). We transformed the variables ‘nPatent’ (OpenMalaria, and OpenMalaria-VECTRI) and ‘PRd’ (VECTRI) into a monthly prevalence. For the VECTRI model, we multiplied the prevalence time series by a scaling coefficient to obtain the same mean between the VECTRI and the OpenMalaria-VECTRI models. We plotted the monthly rainfall to display the rainy seasons. We plotted the VECTRI-modeled emergence rate to show the intermediate step in both the VECTRI and the joint OpenMalaria-VECTRI modelling framework.

**Figure S4:**
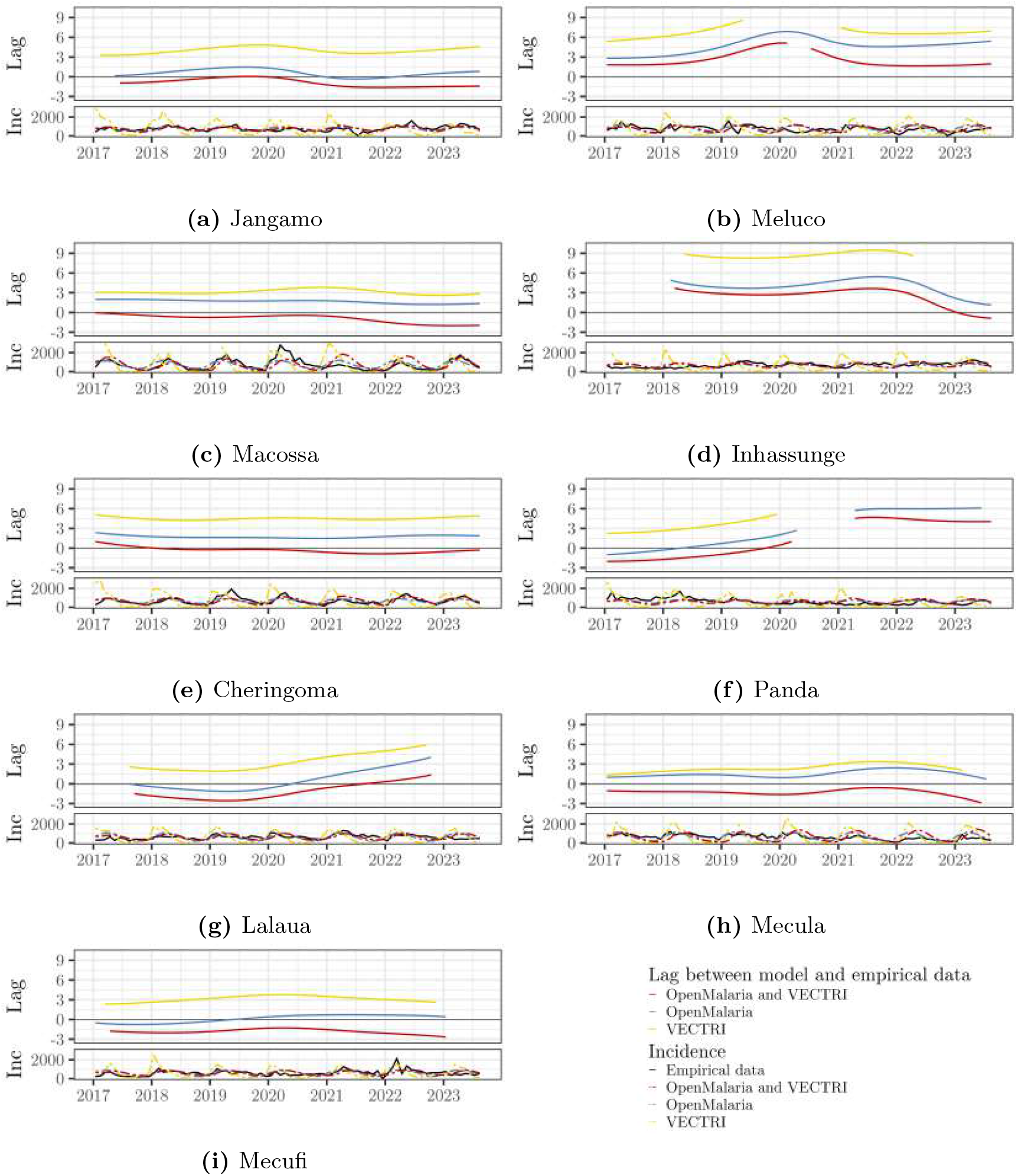
Evolution of the lag between the modeled and the empirical incidence, for the 9 Mozambican districts with the highest incidence after Chibabava (shown in the main article). Lags were obtained through the computation of the phase differences between each model and the empirical data. A positive lag means that the model is ahead of the empirical data. The incidence at the bottom is displayed for reference, and units are per 10,000 people per month.

**Figure S5:**
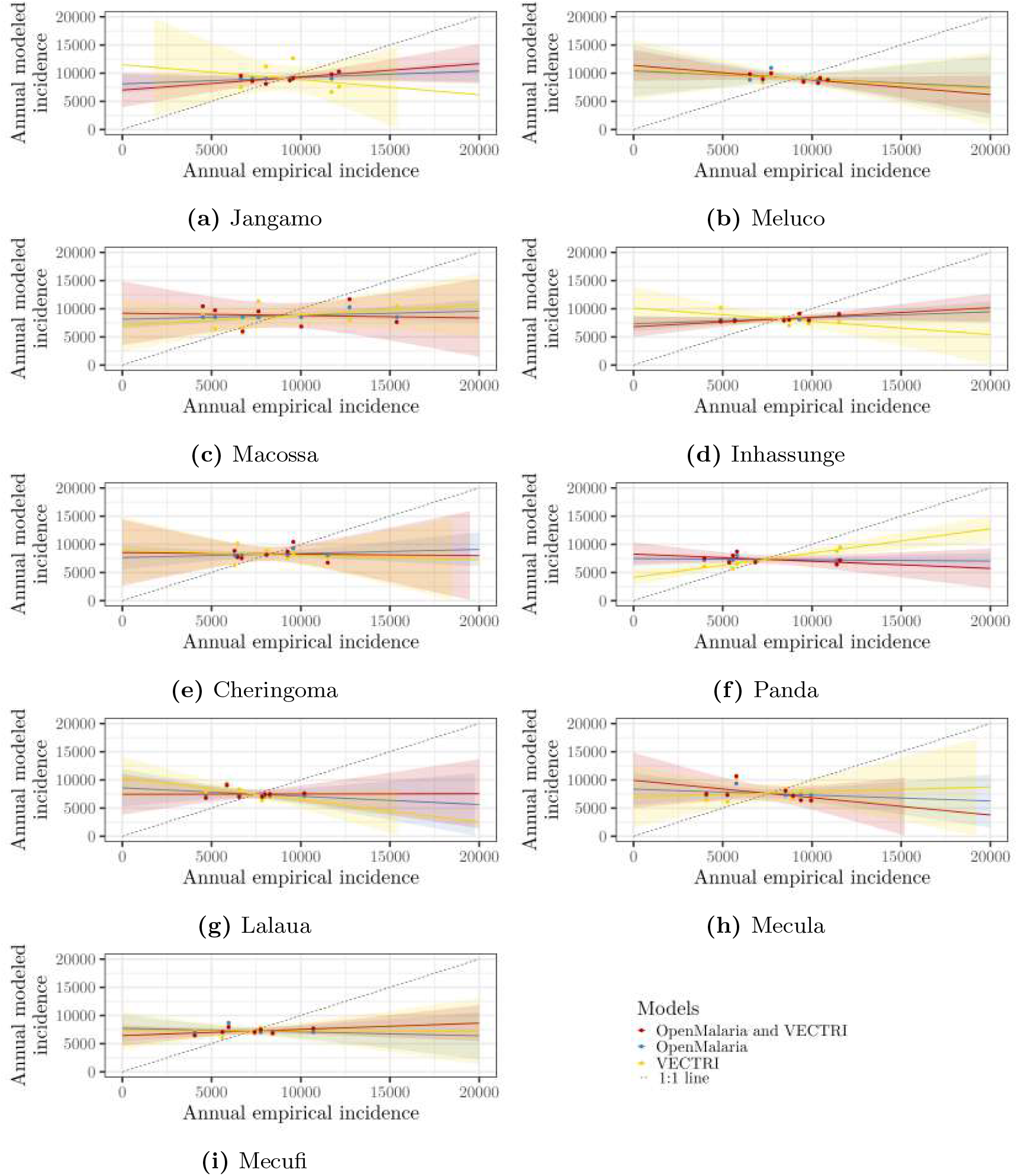
Comparison of the empirical and modeled annual malaria incidence, for the 9 Mozambican districts with the highest incidence after Chibabava (shown in the main article). Each dot represent a year and a model. The x-value is the empirical incidence that year, while the y-value is the predicted incidence for that year, according to the model. For each model, we have scaled the over-all-years modeled incidence to have the same mean as the over-all-years empirical incidence (i.e, we used the time series data from Figure S2). We plotted the 1:1 line for reference.

**Figure S6:**
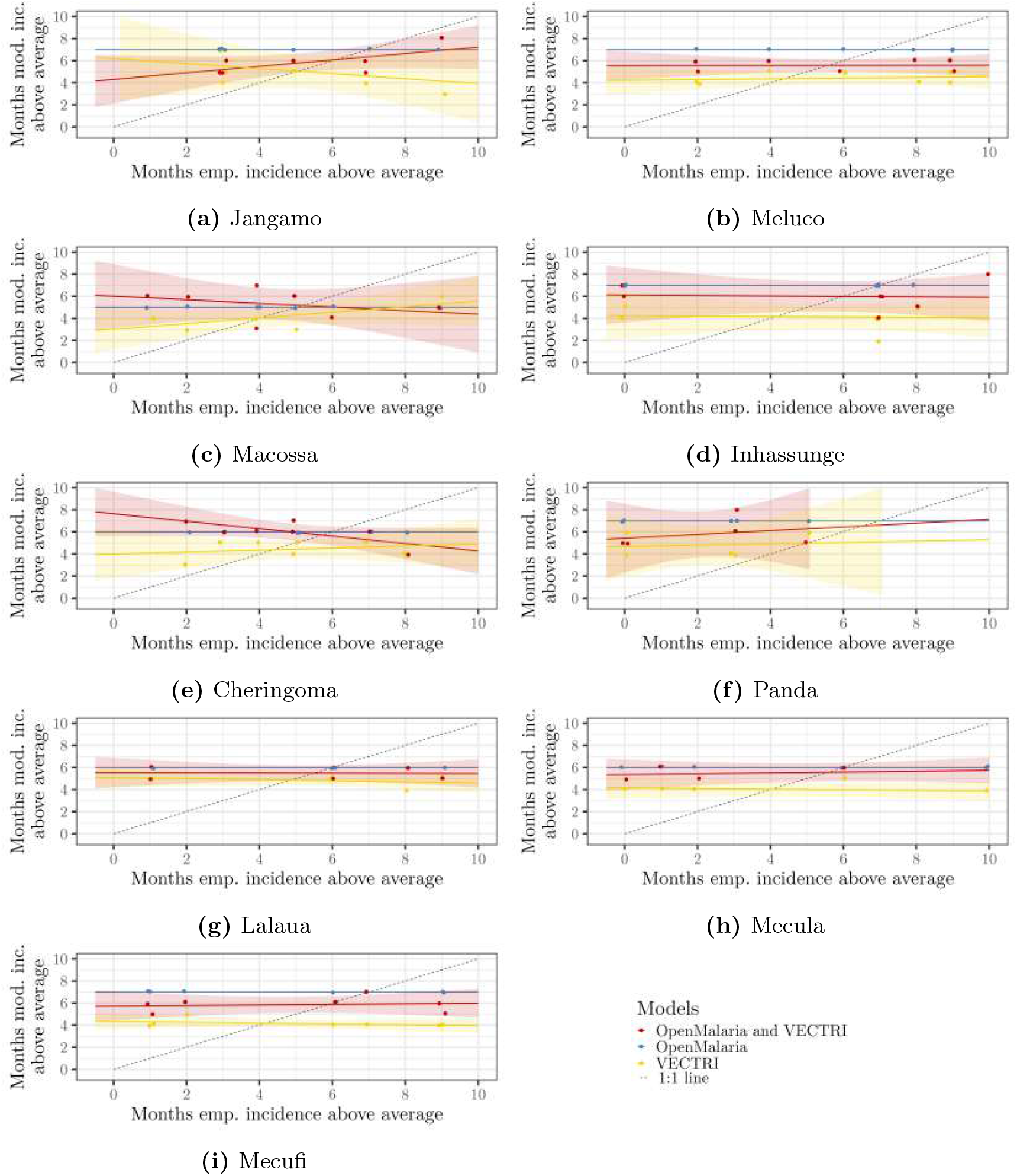
Comparison of the annual number of months with malaria incidence above the average, empirical against predicted by the models, for the 9 Mozambican districts with the highest incidence after Chibabava (shown in the main article). Each dot represent a year and a model. The x-value is the total number of months when the empirical malaria incidence that month was above the all-study-period average. The x-value is the total number of months when the predicted malaria incidence that month was above the all-study-period average, according to the model. For each model, we have scaled the overall modeled incidence to have the same mean as the overall empirical incidence (i.e, we used the time series data from Figure S2). We plotted the 1:1 line for reference.

